# Challenges and perspectives in implementing whole-exome sequencing in Algeria: lessons from a fully autonomous in-country cohort

**DOI:** 10.64898/2026.03.23.26348909

**Authors:** Tarik Ait Mouhoub, Kouider Beladgham, Sihem Brahimi, Nadia Gagi, Ahlam Mihoubi, Hassen Moutchachou, Mohamed El Amine Bouabid, Ahror Belaid, Saïd Yahiaoui, Djamal Belazzougui, Belaid Imessaoudene

**Affiliations:** Laboratory of Metabolic and Rare Genetic Diseases (LMMGR), University of Health Sciences, Algiers, Algeria; Central Biology Laboratory, EHS Ben Aknoun Specialized Hospital, Algiers, Algeria; CERIST, Research Centre for Scientific and Technical Information, Algiers, Algeria

## Abstract

Despite the multidimensional value of implementing genomic medicine, in terms of diagnostic yield, cost-effectiveness, and optimisation of care trajectories, its deployment in many African countries, including Algeria, remains constrained by major structural and interpretive challenges, compounded by the persistent underrepresentation of African populations in genomic databases with direct consequences for variant interpretation and clinical decision-making.

We implemented a fully in-house whole-exome sequencing (WES) workflow structured through a clinically driven sequential framework in 14 unrelated patients with unexplained neurodevelopmental disorders, in a context of high consanguinity and enriched recessive inheritance. A definitive molecular diagnosis was established in 8 cases, with pathogenic or likely pathogenic variants identified in MECP2, PTPN11, FOXG1, ARV1, GNAO1, ATM, ROBO3, and CHD3. Five cases yielded variants of uncertain significance and one clinically relevant incidental finding was identified.

Beyond its diagnostic contribution, this study reveals persistent interpretive limitations: a disproportionate VUS burden, complex incidental finding management, and reduced accessibility to classification criteria, reflecting database underrepresentation, the predominance of private variants, and the limits of current frameworks in consanguineous settings. These findings underscore the necessity of population-specific reference datasets, iterative phenotyping, adapted ethical frameworks, and strategies addressing territorial disparities in access.

This work demonstrates that WES implementation requires a structured multidisciplinary ecosystem integrating clinical, bioinformatic, and ethical dimensions, and provides a transferable model for the sustainable integration of genomic medicine in under-resourced settings, while highlighting the global scientific value of incorporating underrepresented populations into genomic research.

## Introduction

Inequitable access to genomic medicine remains a major challenge across much of Africa, limiting the integration of genetic services into healthcare systems and constraining the development of autonomous diagnostic, analytical, and scientific capacity [1, 2]. This context may also foster inequitable research dynamics, including forms of parachute research, while hindering the emergence of local scientific critical mass and innovation [3, 4]. Prior to this initiative, Algeria, a country of nearly 46 million inhabitants [5], had no alternative but to externalize advanced genomic testing, particularly whole-exome sequencing (WES), thereby addressing only part of the diagnostic needs of rare diseases, 71.9% of which are genetic in origin [6]. This limited access contributes to prolonged diagnostic delay, deferred personalised care, and increased psychological and social burden on affected families, despite the recognized value of early diagnosis-driven management in reducing unnecessary investigations, improving clinical trajectories, and anticipating complications [7]. Whole-exome sequencing has become a first-tier diagnostic tool for neurodevelopmental disorders because of its robust diagnostic performance, clinical utility, and growing capacity to shorten the diagnostic odyssey while guiding personalised management [8,9]. In this context, the implementation of WES for neurodevelopmental disorders represents a strategic priority in Algeria, both for patients and health systems, and for the global scientific community: North African populations, characterised by elevated consanguinity rates and a distinctive recessive variant architecture, remain critically underrepresented in major genomic resources, and this unequal representation directly translates into diagnostic uncertainty and inequity in variant interpretation [10, 11]. However, establishing a WES platform cannot be reduced to its analytical phase alone, as the implementation of a fully autonomous workflow, from clinical expertise to bioinformatic interpretation, requires dedicated medical and bioinformatic expertise, infrastructure, institutional support, and context-specific ethical and interpretive frameworks [1, 2]. By structuring our laboratory around a clinical, technical, and bioinformatic framework relying exclusively on national and public resources, we demonstrate here, for the first time in Algeria, the feasibility and clinical utility of an entirely in-house WES workflow in a first cohort of patients with neurodevelopmental disorders. Beyond its immediate diagnostic contribution, this work provides new insight into the mutational landscape of Algerian patients, offers a concrete model for the development of autonomous genomic medicine programmes in under-resourced settings, and highlights the practical challenges inherent to such implementation, particularly variant interpretation in underrepresented populations, the burden of variants of uncertain significance, infrastructure constraints, and the need for context-specific ethical frameworks.

## Materials and methods (Figure 1)

### Study design, ethics, and consent

This study was conducted in accordance with the principles of the Declaration of Helsinki and received approval from the National Ethics Committee for Health Sciences, Algeria (approval number 2026-02). Written informed consent was obtained from the parents or legal guardians of all paediatric participants, covering diagnostic testing, anonymised research use of clinical and genomic data, and local data storage within Algeria in compliance with national regulations governing the protection of personal and medical data.

**Figure 1.**
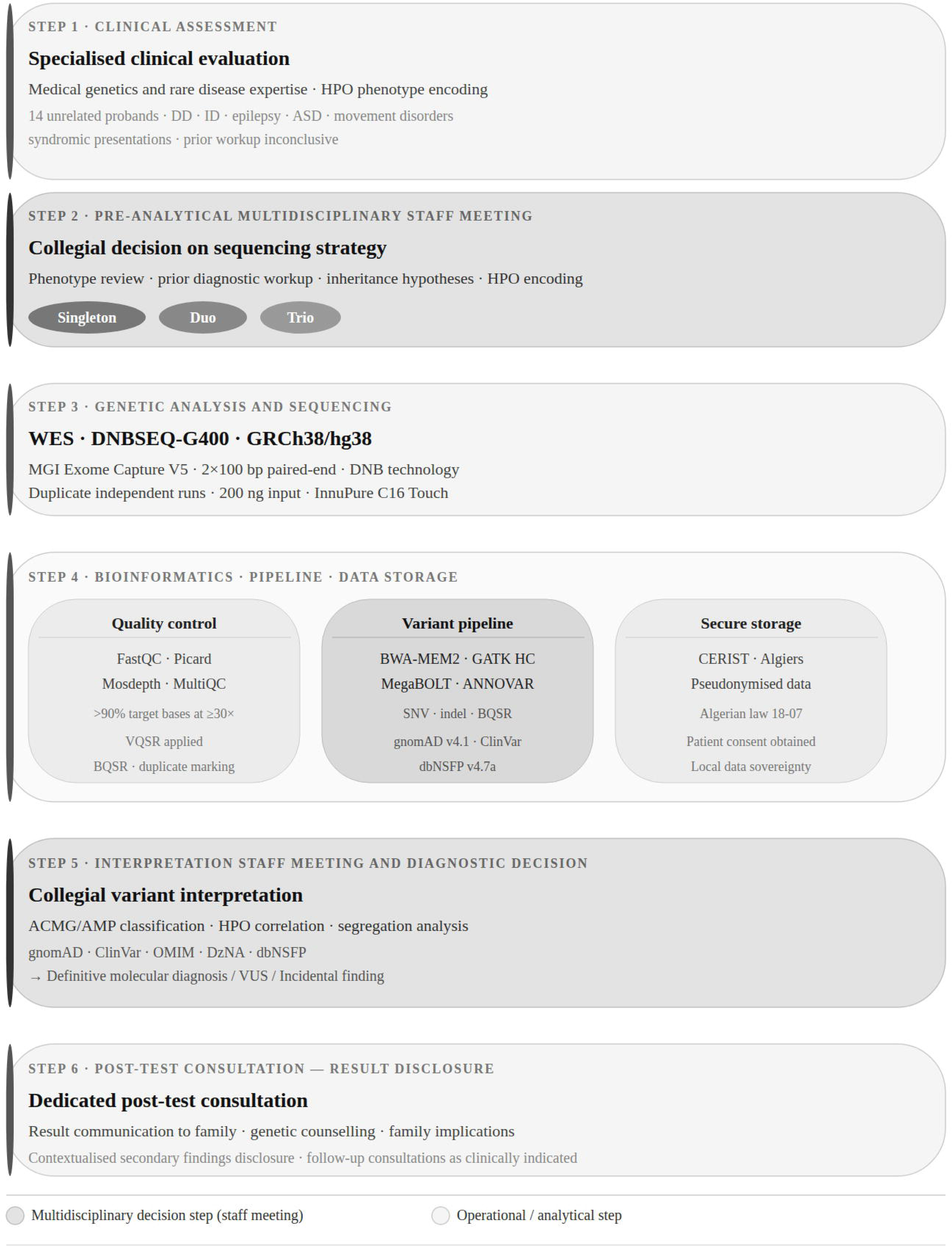
Clinical and bioinformatic workflow for autonomous WES. Each proband underwent specialised clinical evaluation followed by a pre-analytical staff meeting (Step 2) in which phenotype, inheritance hypotheses, and sequencing strategy were collectively agreed. After sequencing and bioinformatic processing, a second staff meeting (Step 5) supported collegial variant interpretation and diagnostic decision. Results were disclosed during a dedicated post-test consultation with provision for follow-up as clinically indicated. ACMG, American College of Medical Genetics and Genomics; AMP, Association for Molecular Pathology; ANNOVAR, Annotate Variation; ASD, autism spectrum disorder; BQSR, base quality score recalibration; BWA-MEM2, Burrows-Wheeler Aligner MEM version 2; ClinVar, clinical variant database (NCBI); DD, developmental delay; DNB, DNA nanoball; dbNSFP, database for nonsynonymous SNP functional predictions; DzNA, Database of Algerian variants and allele frequencies; GATK HC, GATK HaplotypeCaller; gnomAD, Genome Aggregation Database; HPO, Human Phenotype Ontology; ID, intellectual disability; OMIM, Online Mendelian Inheritance in Man; SNV, single nucleotide variant; VUS, variant of uncertain significance; VQSR, variant quality score recalibration; WES, whole-exome sequencing.

### Clinical recruitment, phenotyping, and diagnostic workflow

A total of 14 unrelated probands were referred for genetic evaluation on the basis of unexplained neurodevelopmental disorders encompassing developmental delay, intellectual disability, epilepsy, movement disorders, autism spectrum disorder, and syndromic presentations for which prior clinical and paraclinical investigations had proved inconclusive. Each case first underwent specialised clinical assessment by practitioners with expertise in medical genetics and rare diseases, after which the indication for genetic testing was reviewed within an internal multidisciplinary staff meeting during which the presenting phenotype, prior diagnostic workup, inheritance hypotheses, and most appropriate sequencing strategy were collectively discussed. Depending on clinical context and sample availability, WES was ultimately performed in singleton, duo, or trio configuration. Phenotypic features were encoded using Human Phenotype Ontology (HPO) terms to standardise phenotype capture and facilitate genotype–phenotype correlation [12].

### Sample collection, DNA extraction, and pre-analytical quality assessment

Peripheral blood samples were collected in EDTA tubes from all probands and, whenever available, from both biological parents. Genomic DNA was extracted from peripheral blood leukocytes using the InnuPure C16 Touch automated extraction system (Analytik Jena AG, Jena, Germany) according to the manufacturer’s instructions [13]. DNA purity was evaluated by spectrophotometric measurement of absorbance ratios at OD 260/280 using a Scandrop2 UV/VIS spectrophotometer (Analytik Jena AG, Jena, Germany), with acceptable values defined in the range of 1.8 to 2.0. DNA integrity was verified by agarose gel electrophoresis, and quantification was performed using the Qubit dsDNA HS Assay Kit (Thermo Fisher Scientific, Waltham, MA, USA). Only samples satisfying the quality thresholds established for high-throughput sequencing applications within this laboratory were retained for further processing.

### Library preparation, exome capture, and sequencing

For each sample, 200 ng of input genomic DNA was subjected to library preparation using the MGIEasy FS DNA Library Prep Set (MGI Tech Co., Ltd., Shenzhen, China) according to the manufacturer’s instructions, comprising successive steps of enzymatic fragmentation, end repair, A-tailing, adapter ligation, and pre-amplification. Exome enrichment targeting all coding exons and flanking canonical splice junctions within approximately ±50 bp was performed using the MGIEasy Exome Capture V5 probe set (MGI Tech). For each flow-cell lane, four library pools each comprising eight indexed libraries were prepared; hybridisation with capture probes was carried out in dedicated enrichment buffer, followed by post-hybridisation washing and post-capture PCR amplification. Final libraries were converted into DNA nanoballs, compact concatemers generated by rolling circle amplification of circularised single-stranded DNA templates and constituting the foundational sequencing substrate of DNBSEQ technology [14], before being loaded onto the DNBSEQ-G400 platform (MGI Tech) and sequenced using a paired-end protocol of 2 × 100 bp reads.

To ensure analytical validity and confirm variant calls in the context of first-time platform implementation, the entire sequencing workflow was independently performed on two independently collected blood samples from each individual.

### Primary bioinformatic processing

Primary FASTQ-to-VCF analysis was performed entirely within the local infrastructure using the MegaBOLT bioinformatics hardware accelerator (MGI Tech Co., Ltd., Shenzhen, China), a CPU–FPGA heterogeneous computing platform integrating all primary analysis steps within a single system and achieving speeds substantially exceeding conventional software-based approaches [15]. The pipeline comprised read quality control and filtering, alignment to GRCh38/hg38 using BWA-MEM2 [16], coordinate sorting, PCR duplicate marking, base quality score recalibration (BQSR), and germline variant calling with GATK HaplotypeCaller v3.8, the version embedded within MegaBOLT, following best-practice recommendations [17]. Variant quality score recalibration (VQSR) was applied using standard SNV and indel filtration thresholds. Sequencing quality metrics were assessed with FastQC and aggregated via MultiQC; alignment quality, including mean target coverage, fold enrichment, and breadth of coverage, was evaluated using Picard CollectHsMetrics and independently validated with Mosdepth against a BED file of RefSeq-annotated coding exons and canonical splice sites. This workflow consistently produced high-quality variant call files, with greater than 90% of target bases achieving a minimum sequencing depth of 30×.

### Secondary annotation and variant interpretation

Secondary analysis used an in-house ANNOVAR-based annotation pipeline integrating gnomAD v4.1 exome (downloaded 1 September 2025), ClinVar release 20240917, dbSNP build 151, UCSC cytoband annotation, dbNSFP v4.7a, and OMIM gene–disease data. Retained annotation fields encompassed chromosomal coordinates, gene symbol, functional consequence, Human Genome Variation Society (HGVS) protein-level change [20], gnomAD allele frequency, ClinVar classification, OMIM relationships, gene-level constraint metrics, and in silico pathogenicity scores (SIFT4G, MutationTaster, MetaRNN, REVEL, MVP, MPC, AlphaMissense, CADD, GERP++ RS), supplemented by InterPro domain annotations and GTEx v8 eQTL data. Variants were additionally cross-referenced against DzNA (Database of Algerian variants and allele frequencies), an in-house North African population reference developed to mitigate VUS burden from database underrepresentation. Classification followed ACMG/AMP guidelines [18] with explicit application of ClinGen computational evidence criteria [19]; reference transcripts are specified in the variant table.

### Segregation analysis and variant selection

Family-based analysis was conducted in duo or trio configuration when parental DNA was available, to determine inheritance mode and establish or exclude de novo occurrence; when parental samples were unavailable, interpretation relied on phenotype concordance, gene–disease association strength, and published pathogenicity evidence, with the absence of segregation data explicitly documented. Only clinically relevant variants, selected scientifically informative VUS, and incidental findings requiring contextual interpretation were retained for this report.

## Results

Among the 14 cases reported here (Table 1), eight received a definitive molecular diagnosis through the identification of pathogenic or likely pathogenic variants, five were assigned a variant of uncertain significance (VUS) as the primary interpretive outcome, and one yielded an incidental finding of clinical relevance unrelated to the presenting phenotype (Table 2 and 3).

### Definitive molecular diagnoses

#### Previously established pathogenic or likely pathogenic variants with strong clinicogenetic concordance

Five cases corresponded to previously established pathogenic variants showing strong clinicogenetic concordance and allowing straightforward diagnostic attribution (Tables 1, 2 and 3).

##### MECP2

In Patient 2, a girl with a classical Rett syndrome (RTT; MIM #312750) [21] phenotype, including developmental regression, stereotypies, loss of speech, seizures, scoliosis, and breathing dysregulation, we identified a de novo hemizygous MECP2 variant (NM_001110792.2: c.334C>G; p.Leu112Val), absent from gnomAD and previously reported as pathogenic in ClinVar, supporting a definitive diagnosis despite the absence of formal de novo confirmation in singleton context.

PTPN11. WES in Patient 5, whose phenotype was highly suggestive of Noonan syndrome (NS1; MIM #163950) [22], identified a de novo heterozygous PTPN11 variant (NM_002834.5: c.1507G>A; p.Gly503Arg), also absent from gnomAD and previously established as pathogenic, fully consistent with the clinical presentation.

##### FOXG1

In Patient 7, a girl with developmental delay, seizures, and hypotonia, a heterozygous nonsense FOXG1 variant (NM_005249.5: c.654C>A; p.Tyr218Ter) was identified; its predicted loss-of-function effect and the strong phenotype concordance supported classification as pathogenic, consistent with FOXG1 syndrome (MIM #613454) [23].

##### ARV1

In Patient 4, a girl born to consanguineous parents with severe developmental and epileptic encephalopathy, microcephaly, absent speech, autistic behaviour, and elevated alkaline phosphatase, we identified a homozygous frameshift ARV1 variant (NM_022786.3: c.510dupA; p.Pro174AlafsTer14), previously reported as pathogenic and fully concordant with ARV1-related developmental and epileptic encephalopathy (DEE38; MIM #617020) [24].

##### GNAO1

Finally, in Patient 13, a girl with severe early-onset hyperkinetic movement disorder and developmental delay, a heterozygous GNAO1 variant (NM_020988.3: c.736G>A; p.Glu246Lys) was identified; the variant was absent from gnomAD, previously reported as pathogenic in ClinVar, and occurred de novo, supporting a definitive molecular diagnosis and classification as pathogenic, consistent with GNAO1-related neurodevelopmental disorder with involuntary movements (NEDIM; MIM #617493) [25]. Collectively, these cases illustrate situations in which WES rapidly confirmed a clinically coherent diagnosis through variants already well established in the literature and clinical databases.

#### Strong clinicogenetic diagnoses without prior ClinVar representation

Three cases yielded likely pathogenic variants without prior ClinVar representation, for which the diagnostic interpretation relied on the convergence of the identified variant, a predicted effect consistent with the known pathogenic mechanism of the gene, and a highly specific or well-delineated phenotype (Tables 1, 2 and 3).

##### ATM

Trio analysis in Patient 3, a girl born to consanguineous parents and presenting with psychomotor delay, dysarthria, akinesia, abnormal conjunctival vasculature, cafe-au-lait spots, and recurrent respiratory infections, identified a homozygous frameshift variant in ATM (NM_000051.4: c.1951_1952insTA; p.Phe651TyrfsTer13), inherited from both parents, absent from gnomAD, and introducing a premature stop codon in a gene for which biallelic loss of function is an established pathogenic mechanism. The phenotypic concordance with ataxia-telangiectasia (AT; MIM #208900) further supported classification as likely pathogenic (PVS1, PM2, PP4), with direct implications for familial cancer-risk counselling.

##### ROBO3

In Patient 10, a boy born to consanguineous parents, WES identified a homozygous frameshift variant in ROBO3 (NM_022370.4: c.2579_2580del; p.Val860AlafsTer125), inherited from both parents, absent from gnomAD, and predicted to result in premature truncation, supporting classification as likely pathogenic (PVS1, PM2). The clinical findings of gaze-evoked horizontal nystagmus, progressive congenital scoliosis, hypotonia, pontocerebellar hypoplasia on brain MRI, and a family history of similarly affected siblings were fully concordant with horizontal gaze palsy with progressive scoliosis (HGPPS; MIM #607313) [26], thereby establishing the diagnosis through WES.

##### CHD3

A heterozygous frameshift variant in CHD3 (NM_001005273.3: c.9delG; p.Ala4GlnfsTer4), absent from gnomAD and predicted to result in loss of function in a gene showing strong loss-of-function constraint (oe_lof = 0.20), fully concordant with Snijders Blok-Campeau syndrome (SBCS; MIM #618205) [27], was identified in Patient 12, a girl presenting with intellectual disability, global developmental delay, postnatal macrocephaly, dystonia, and hypomyelination with callosal dysgenesis on brain MRI. Parental samples were unavailable, precluding formal de novo confirmation, but the overall evidence supported classification as likely pathogenic (PVS1, PM2).

### Variants of uncertain significance

#### VUS in genes with plausible neurodevelopmental relevance but insufficient evidence (Tables 1, 2 and 3)

##### SMG8

Trio analysis in Patient 1, a boy with severe syndromic intellectual disability born to a first-cousin couple, identified a homozygous missense variant in SMG8 (NM_018149.7: c.2668T>C; p.Tyr890His), inherited from both parents. The variant has been observed only once in gnomAD in the homozygous state, in an individual classified under the ‘remaining ancestry’ category, and once in ClinVar (Variation ID: 4170574) as a variant of uncertain significance. The phenotypic spectrum of Alzahrani-Kuwahara syndrome (ALKUS; MIM #619268) was initially described with non-constant features including microcephaly and ocular malformations [28]; subsequent reports indicated that deepening of nasolabial folds with age represents the most constant sign, a feature not observed in our patient [29]. The observation of more severe phenotypes with truncating variants raises the question of a hypomorphic versus null-allele effect [28,30], counterbalanced here by the severity of the presentation despite a missense mutation. In the absence of additional segregations or functional studies, the variant was classified as VUS (PM2).

##### ARFGEF1

A heterozygous missense variant in ARFGEF1 (NM_006421.5: c.622C>T; p.Arg208Cys) was identified in Patient 14, a boy with early developmental impairment; the variant was absent in the mother, while the father was unavailable for testing. ARFGEF1 haploinsufficiency has been associated with developmental delay and epilepsy with incomplete penetrance and variable expressivity (MIM #619964) [31], and this gene displays strong overall intolerance to missense variation (mis_z = 6.96); however, the genomic constraint analysis of the surrounding 1 kb region shows a local z-score of 0.17, which is not statistically significant. The variant is extremely rare in gnomAD (AF 8.21 x 10-6) and has not been reported in ClinVar. In silico scores are predominantly in favour of pathogenicity, but the overall weight of this argument remains limited. In the absence of segregation data, functional studies, or independent cases carrying the same variant, the variant was classified as VUS (BP1, PP3).

##### MED12L

MED12L pathogenic variants cause Nizon-Isidor syndrome (NIZOS; MIM #618872), a spectrum recently expanded to include white matter abnormalities [32] and maternal inheritance [33]. In Patient 11, a boy with syndromic intellectual disability born to consanguineous parents, a heterozygous missense variant in MED12L (NM_053002.6: c.2365A>G; p.Lys789Glu) was identified. Parental samples were unavailable for segregation analysis. Although the missense constraint score for this gene exceeds 4, the local 1 kb constraint z-score (gnomAD, z = 1.13) was not significant. The variant was classified as VUS (PM2, PP3, BP1).

#### VUS limited by unresolved gene–disease validity or insufficient specificity (Tables 1, 2 and 3)

##### UPB1

Although biallelic UPB1 variants have been associated with a neurodevelopmental phenotype (MIM #613161) [34] and functional loss of enzymatic activity has been demonstrated [35], a neonatal screening study identified ten children with biochemical beta-ureidopropionase deficiency of whom nine remained clinically asymptomatic [36], leaving the causal link to neurological disease unestablished. In Patient 8, a girl with severe syndromic intellectual disability born to consanguineous parents, a homozygous missense variant in UPB1 (NM_016327.3: c.319G>A; p.Ala107Thr) was identified. No specific biochemical investigations were performed. The variant was classified as VUS (PM2).

##### ZC3H14

A homozygous missense variant in ZC3H14 (NM_001326314.2: c.566T>A; p.Leu189His), absent from gnomAD and predicted to be deleterious by multiple in silico tools, was identified in Patient 9, a boy with severe intellectual disability born to non-consanguineous parents. Truncating variants in ZC3H14 have been reported in two consanguineous families with non-syndromic intellectual disability (MRT56; MIM #617125) with functional support [37], but the gene-disease relationship remains of limited evidence validity and ClinVar submissions reporting pathogenicity (Variation ID: 254278) are flagged with reason of insufficient supporting evidence. In the absence of segregation data and functional validation, the variant was classified as VUS (PM2, PP3).

#### Incidental finding and interpretive implications

##### RYR1

In Patient 6, a girl with severe syndromic intellectual disability born to consanguineous parents, a heterozygous variant in RYR1 (NM_000540.3: c.10579C>T; p.Pro3527Ser) was identified and inherited from the father, who reported no personal or familial history of malignant hyperthermia or anaesthetic complications. The variant was extremely rare in gnomAD and is classified in ClinVar as pathogenic for autosomal recessive congenital multicore myopathy (CMYO1B; MIM #255320), based on prior reports in a consanguineous Algerian family [38]. In the present context, this finding was consistent with carrier status and did not explain the presenting phenotype. The critical question raised was whether this variant could confer malignant hyperthermia (MH) susceptibility; the patient’s features did not correspond to King-Denborough syndrome (KDS; MIM #619542), the variant has not been reported as causative for MH, and no functional studies have addressed this question. In keeping with international recommendations on secondary findings [39] and RYR1-specific classification frameworks, we could not formally implicate this variant in MH susceptibility (Tables 1, 2 and 3).

## Discussion

Beyond diagnostic yield, our data highlight that the implementation of WES in underrepresented populations fundamentally reshapes the interpretive, ethical, and organisational dimensions of genomic medicine.

This study confirms the clinical contribution of WES, enabling the end of a long diagnostic odyssey (Patients 2, 5, 11) and the establishment of early diagnosis (Patient 13). The corollary is better personalised management, the adequation of follow-up and anticipatory strategies, the limitation of unnecessary paraclinical investigations, and the reduction of the psychological burden of this diagnostic wandering for both families and patients [40]. The gain in terms of cost also represents a significant argument for the integration of this type of workflow into a healthcare system in African countries [40].

A further dimension of this work, which underlines the necessity of establishing a contextual clinical and ethical approach adapted to the local landscape, concerns incidental findings. The RYR1 variant identified in Patient 6 was detected in the heterozygous state and does not account for the presenting neurodevelopmental phenotype. The clinical question it raises, namely potential susceptibility to malignant hyperthermia, required interpretation according to gene-specific classification frameworks, in the absence of directly applicable functional data. Although international recommendations exist regarding the management of secondary findings [39], their application cannot be mechanically transposed without adaptation to local anthropological, cultural, and systemic contexts [41]. A contextually anchored approach to secondary findings, integrating family consent, local ethics frameworks, and culturally appropriate conditions for result disclosure, is therefore essential and constitutes ongoing work within the laboratory.

Similarly, the identification of a biallelic ATM frameshift variant in Patient 3, consistent with a diagnosis of ataxia-telangiectasia, raises the clinically critical issue of managing heterozygous carrier status in the parents and at-risk family members. Heterozygous germline pathogenic variants in ATM are associated with an increased risk of several cancers, including female breast cancer [42], with a moderately increased risk for pancreatic cancer [43] and, for male carriers, a potential increased susceptibility to prostate cancer [44]. Penetrance remains incomplete and expressivity variable, particularly for previously unreported variants [45].

In the present family, no personal or family history of cancer was reported at the time of consultation. Current international guidelines recommend individualised risk assessment, incorporating the type of variant, personal and family oncological history, age, and sex. For carriers of truncating or equivalent-risk variants without a personal cancer history, breast cancer surveillance is indicated, with modalities and initiation age adapted to estimated lifetime risk and applicable national guidelines [42,45]; prostate surveillance may be considered according to applicable guidelines and individual risk assessment [44]; pancreatic surveillance is recommended only within a research framework [43]. The disclosure of these results to the parents, as obligate heterozygous carriers, therefore requires a structured oncogenetic consultation, adapted to the local context, and integrated within a multidisciplinary decision-making process that explicitly incorporates family consent and culturally appropriate conditions for result disclosure [41]. This case exemplifies how the identification of a recessive paediatric condition through WES can open an unsolicited oncogenetic dimension for parents and underscores the necessity of an organised, contextualised oncogenetics framework as a companion to any clinical WES programme.

Our study also highlights a major issue in underrepresented populations in databases, making certain ACMG classification criteria less accessible, namely the burden of VUS. Although WES diagnostic yield in neurodevelopmental disorders is broadly comparable across clinical indications regardless of genetic ancestry [46], variant interpretation remains substantially more challenging in underrepresented populations, due to a higher proportion of rare or previously unreported variants in reference databases such as gnomAD and ClinVar, resulting in a disproportionate VUS burden [5,10]. This cohort represents only a preliminary sample of our structure, but one must expect that the frequency of variants absent from public databases creates a disproportionate burden of VUS classification and diagnostic uncertainty, highlighting the urgent need for population-specific reference databases and data-sharing initiatives to improve ACMG-based variant classification [47].

Our data further reveal that several VUS cases involve genes with limited gene–disease validity (ZC3H14, UPB1), where the question is not merely one of variant classification but of the underlying gene–disease relationship itself. These observations underscore the need for regular bioinformatic reanalysis [48] and for functional validation studies, resources that remain difficult to mobilise in settings where genomic research capacity is still being built. More generally, they also raise the question of refining phenotype. Indeed, the apparent clinical discordance may reflect either true variability (genetic modifiers, consanguinity, environment), or an incomplete phenotype prior to reverse phenotyping. Moreover, the phenotypic variability of monogenic diseases has been partially constructed on predominantly European cohorts, limiting transferability and biasing interpretation [5,49], whereas enrichment in underrepresented populations may allow expansion of phenotypic spectra, better estimation of penetrance, and reduction of blind spots in precision medicine.

The challenges raised by our approach operate at two interconnected levels: interpretive challenges, including population underrepresentation in genomic databases, a disproportionate VUS burden, reduced accessibility of ACMG criteria in the context of consanguinity and recessive inheritance, eurocentrism of established phenotypic spectra, and the need for anthropologically contextualised ethical frameworks and structural challenges, encompassing reagent supply constraints, sustainable funding, laboratory accreditation, the development of a local genomic scientific culture, integration into a transdisciplinary clinical workflow, and the risk of territorial disparities in access [47,50].

To address these challenges, a transversal national approach is required. Our decision to structure this workflow according to a sequential clinical–technical–bioinformatic–interpretative clinical approach establishes from the outset a strict framework, avoiding reduction of WES to a simple sequencing act, given the complexity of interpretation and associated ethical risks. In this perspective, the development of locally adapted ethical guidelines, training programmes in medical genetics and genetic counseling, and national clinical and scientific networks appears essential, as ethical frameworks cannot be directly transposed between countries and must be anchored in local cultural, social, and healthcare systems [41].

In order to reduce territorial disparities and refine phenotype enrichment based on HPO coding [12], a national application for clinical genetic referral has been developed (AMINgen: Application for Medical Inquiry and Nexus in Genomics), enabling a tight clinician–geneticist–patient interface and anchoring this practice within a structured and integrated clinical framework. Furthermore, the DzNA database affiliated with our laboratory constitutes both a variant database and a population reference database, providing two complementary entry axes that will progressively improve variant interpretation as its implementation expands [47,49]. The establishment of a learned society structured around technical (workflow standardisation and validation), ethical (indications, VUS, incidental findings, integration of patient and family perspectives, anthropological contextualisation), and academic (training, genetic counseling, dissemination) dimensions, in coordination with trans-ministerial governance (funding, logistics, accreditation, maintenance, regulatory structuring), appears necessary to ensure the sustainability, sovereignty, and secure clinical integration of genomic medicine for the benefit of patients [41,50] (Figure 2).

**Figure 2.**
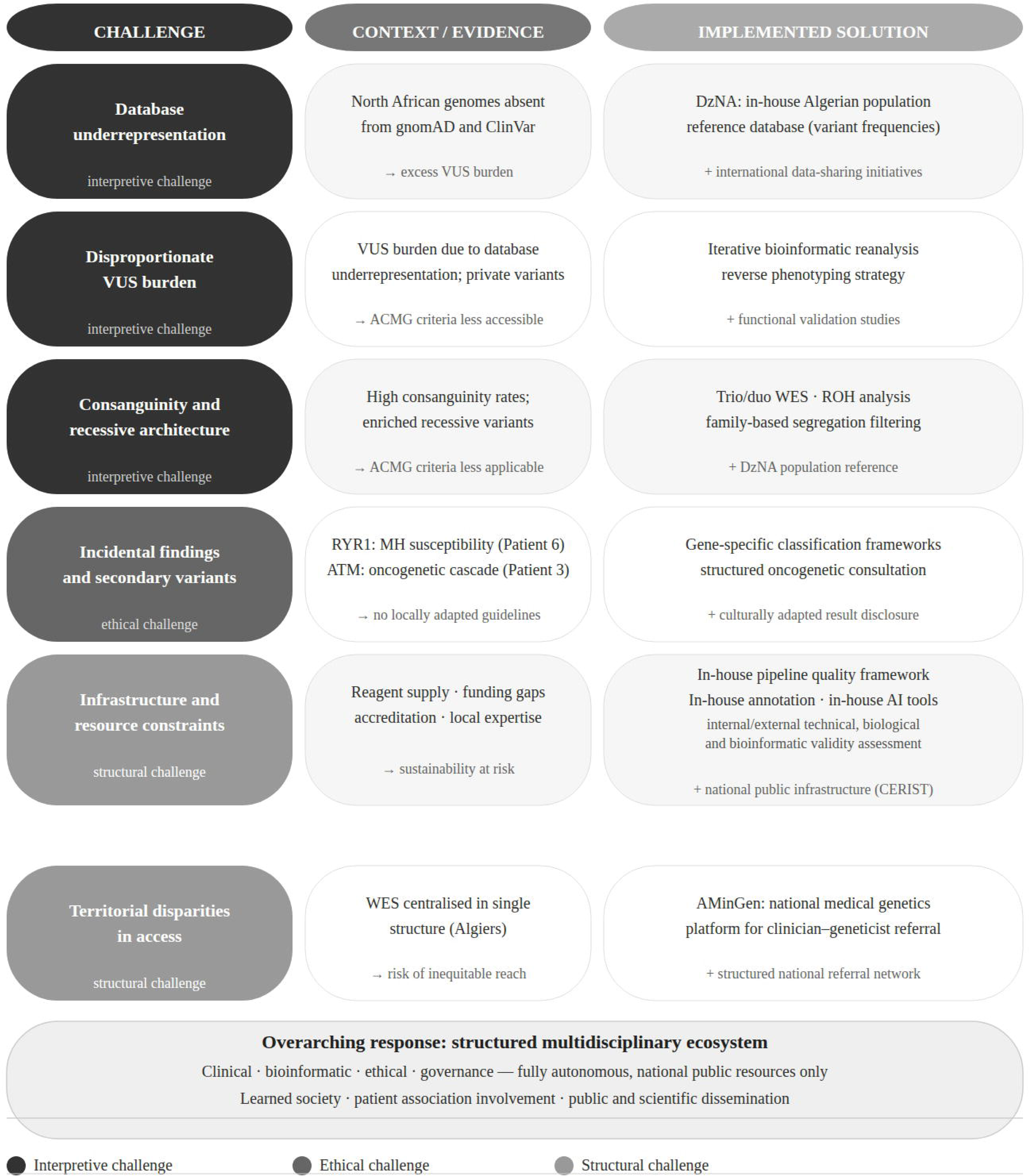
Key challenges encountered during autonomous WES implementation in Algeria, with contextual evidence and implemented or planned solutions. Challenges operate at two interconnected levels: interpretive (dark grey, relating to population underrepresentation, VUS burden, and consanguinity) and ethical/structural (medium grey, relating to incidental findings, infrastructure, and territorial access). Each challenge is paired with its evidentiary context and the corresponding response developed within this programme. ACMG, American College of Medical Genetics and Genomics; AI, artificial intelligence (here: in-house computational tools for variant prioritisation and interpretation); AMinGen, Application for Medical Inquiry and Nexus in Genornics (national clinical genetics referral platform); AMP, Association for Molecular Pathology; ATM, ataxia telangiectasia mutated gene; CERIST, ResearchCentre for Scientific and Technical Information; DzNA, Database of Algerian variants and allele frequencies; HPO, Human Phenotype Ontology; MH, malignant hyperthermia; ROH, runs of homozygosity; RYRl, ryanodine receptor 1 gene; VUS, variant of uncertain significance; WES, whole-exome sequencing.

Finally, we have to note that our cohort size is limited and reflects a first national experience within a single structure, restricting generalisability. The absence of functional validation studies and limited longitudinal follow-up reduce our capacity to fully characterise genotype–phenotype correlations and penetrance for emerging gene–disease associations. Furthermore, although the DzNA national variant database has been established, its current limited size precludes any substantial mitigation of the population underrepresentation bias affecting variant classification, and its progressive enrichment will be essential to improve the interpretive yield of future cohorts.

In conclusion, this study establishes the clinical and scientific feasibility of autonomous whole-exome sequencing within a public health framework in Algeria, and underscores that the challenges identified cannot be addressed through the mechanical transposition of frameworks developed in high-income settings, but require multidimensional, locally anchored solutions that reflect the specific genomic, cultural, and institutional landscape of each country. These findings offer a replicable model for the development of sovereign genomic medicine programmes across similarly positioned settings.

## Data availability

All data are stored and secured at the Laboratory of Metabolic and Rare Genetic Diseases (LMMGR) and the Research Centre for Scientific and Technical Information (CERIST), Algiers, under Algerian law 18-07 and are available from the corresponding author upon reasonable requestand subject to applicable national regulations.

## Supporting information

Table 2: Genetic findings and variant annotation

Table 3: In silico prediction scores and functional annotations

Table 1: Clinical characteristics of the cohort

## Acknowledgements

The authors thank the Ministry of Higher Education and Scientific Research of Algeria for institutional support, and express their deepest gratitude to the patients and their families for their participation and trust.

## Author information

Authors and Affiliations

**Laboratory of Metabolic and Rare Genetic Diseases (LMMGR), University of Health Sciences, Algiers, Algeria**

Tarik AIT MOUHOUB, Kouider BELADGHAM, Sihem BRAHIMI, Nadia GAGI, Ahlam MIHOUBI and Belaid IMESSAOUDENE

**Central Biology Laboratory, EHS Ben Aknoun Specialized Hospital, Algiers, Algeria**

Sihem BRAHIMI, Nadia GAGI, Ahlam MIHOUBI and Belaid IMESSAOUDENE

**CERIST, Research Centre for Scientific and Technical Information, Algiers, Algeria**

Djamal BELAZZOUGUI, Saïd YAHIAOUI, Hassen MOUTCHACHOU, Mohamed El Amine BOUABID and Ahror BELAID

## Contributions

TAM, KB and BI conceptualised the study provided overall supervision and project administration. TAM, DB, SY, HM, MEAB and AB designed and implemented the bioinformatic pipeline. TAM, KB and SB performed clinical assessment and variant interpretation. NG and AM performed sequencing and contributed to variant interpretation. DB, SY, HM, MEAB and AB developed the computational infrastructure and contributed to bioinformatic analysis. TAM and KB wrote the manuscript. All authors reviewed and edited the manuscript.

## Corresponding author

Correspondence to Tarik AIT MOUHOUB

## Ethics declarations

### Ethical approval and consent to participate

This study was conducted in accordance with the principles of the Declaration of Helsinki and received approval from the National Ethics Committee for Health Sciences, Algeria (approval number 2026-02). Written informed consent was obtained from the parents or legal guardians of all pediatric participants.

