## Supplementary material for "Challenges and perspectives in implementing whole-exome sequencing in Algeria: lessons from a fully autonomous in-country cohort": Table 2: Genetic findings and variant annotation

### Table 2. Genomic findings identified by whole-exome sequencing.

*Pathogenic (P), likely pathogenic (LP), and variants of uncertain significance (VUS) identified across the 14 patients, together with the incidental finding (Patient 6, RYR1). ACMG/AMP classification criteria are listed in parentheses. Genomic position refers to GRCh38/hg38 coordinates. Inheritance is based on parental segregation when available; — indicates absent or not reported.*

| Pt | Gene | Transcript | Variant (cDNA(protein)) | Zyg. | Inheritance | ACMG class (criteria) | WES config. | Genomic position | Depth | Reads Ref | Reads Alt | avsnp151 | WES outcome |
|---|---|---|---|---|---|---|---|---|---|---|---|---|---|
| **1** | **SMG8** | NM_018149.7 | c.2668T>C(p.Tyr890His) | Hom | Inherited (both parents) | VUS (PM2) | Trio | 59213491 | 53 | 2 | 51 | rs137980220 | VUS |
| **2** | **MECP2** | NM_001110792.2 | c.334C>G(p.Leu112Val) | Hem | De novo | P (PP5, PM1, PP3, PM5, PM2, PS2) | Singleton | 154032286 | 38 | 17 | 21 | rs28935168 | Diagnostic |
| **3** | **ATM** | NM_000051.4 | c.1951_1952insTA(p.Phe651TyrfsTer13) | Hom | Inherited (both parents) | LP (PVS1, PM2, PP4) | Trio | 108253866 | 82 | 1 | 81 | — | Diagnostic |
| **4** | **ARV1** | NM_022786.3 | c.510dupA(p.Pro174AlafsTer14) | Hom | Inherited (both parents) | P (PVS1, PM2, PP5) | Trio | 230995820 | 37 | 5 | 32 | rs544784472 | Diagnostic |
| **5** | **PTPN11** | NM_002834.5 | c.1507G>A(p.Gly503Arg) | Het | De novo | P (PP5, PS1, PM1, PM5, PP3, PM2, PS2, PP4) | Trio | 112489083 | 28 | 14 | 14 | rs397507545 | Diagnostic |
| **6** | **RYR1** | NM_000540.3 | c.10579C>T(p.Pro3527Ser) | Het | Inherited (father) | P (PP2, PP5) | Trio | 38525455 | 31 | 17 | 14 | rs118192164 | Incidental finding |
| **7** | **FOXG1** | NM_005249.5 | c.654C>A(p.Tyr218Ter) | Het | Assumed de novo | P (PVS1, PM2, PS4, PP4) | Singleton | 28767933 | 28 | 11 | 17 | — | Diagnostic |
| **8** | **UPB1** | NM_016327.3 | c.319G>A(p.Ala107Thr) | Hom | Inherited (both parents) | VUS (PM2) | Singleton | 24502168 | 132 | 3 | 129 | rs770926451 | VUS |
| **9** | **ZC3H14** | NM_001326314.2 | c.566T>A(p.Leu189His) | Hom | Inherited (both parents) | VUS (PM2, PP3) | Singleton | 88575848 | 159 | 0 | 159 | — | VUS |
| **10** | **ROBO3** | NM_022370.4 | c.2579_2580del(p.Val860AlafsTer125) | Hom | Inherited (both parents) | LP (PVS1, PM2) | Singleton | 124876110 | 21 | 0 | 21 | — | Diagnostic |
| **11** | **MED12L** | NM_053002.6 | c.2365A>G(p.Lys789Glu) | Het | Parents unavailable | VUS (PM2, PP3, BP1) | Singleton | 151355192 | 105 | 53 | 52 | — | VUS |

| | | | | | | | | | | | | |
|---|---|---|---|---|---|---|---|---|---|---|---|---|
| **12** | **CHD3** | NM_001005273.3 | c.9delG(p.Ala4GlnfsTer4) | Het | Parents unavailable | LP (PVS1, PM2) | Singleton | 7889009 | 30 | 14 | 16 | — | Diagnostic |
| **13** | **GNAO1** | NM_020988.3 | c.736G>A(p.Glu246Lys) | Het | De novo | P (PP5, PM5, PM1, PP3, PS2) | Singleton | 56351396 | 34 | 17 | 17 | rs797044951 | Diagnostic |
| **14** | **ARFGEF1** | NM_006421.5 | c.622C>T(p.Arg208Cys) | Het | Mother negative; father unavailable | VUS (BP1, PP3) | Duo | 67296448 | 109 | 57 | 52 | rs763667176 | VUS |

*ACMG, American College of Medical Genetics and Genomics; AMP, Association for Molecular Pathology; avsnp151, dbSNP build 151 identifier; Hem, hemizygous; Het, heterozygous; Hom, homozygous; LP, likely pathogenic; mo, months; P, pathogenic; Pt, patient; VUS, variant of uncertain significance; WES, whole-exome sequencing; Zyg., zygosity.*
