## Supplementary material for "Challenges and perspectives in implementing whole-exome sequencing in Algeria: lessons from a fully autonomous in-country cohort": Table 3: In silico prediction scores and functional annotations

**Table 3. In silico pathogenicity prediction scores and population frequency data.**

*In silico pathogenicity scores for all variants identified in the study cohort. gnomAD allele frequencies are from the gnomAD v4.1 exome dataset; Absent = not observed in gnomAD. Scores of — indicate non-applicable (frameshift, nonsense) or unavailable values. oe_lof and mis_z are gene-level constraint metrics from gnomAD v4.1; lower oe_lof and higher mis_z indicate stronger intolerance to loss-of-function and missense variation, respectively.*

| Patient | Gene | Variant (cDNA(protein)) | gnomAD AF (v4.1) | ClinVar | SIFT4G | Mutation Taster | MetaRNN | REVEL | MVP | MPC | Alpha Missense | CADD (phred) | GERP++ RS | oe_lof (v4.1) | mis_z (v4.1) |
|---|---|---|---|---|---|---|---|---|---|---|---|---|---|---|---|
| **1** | **SMG8** | c.2668T>C(p.Tyr890His) | 0.0002 | VUS | 0.000 | 1.000 | 0.089 | 0.638 | 0.623 | 0.634 | 0.703 | 26.5 | 5.85 | 0.567 | 2.865 |
| **2** | **MECP2** | c.334C>G(p.Leu112Val) | Absent | Pathogenic | 0.197 | 1.000 | 0.967 | 0.797 | 0.999 | — | 0.982 | 24.2 | 5.06 | — | — |
| **3** | **ATM** | c.1951_1952insTA(p.Phe651TyrfsTer13) | Absent | — | — | — | — | — | — | — | — | — | — | 0.767 | 2.541 |
| **4** | **ARV1** | c.510dupA(p.Pro174AlafsTer14) | 6.71e-05 | Pathogenic | — | — | — | — | — | — | — | — | — | 0.804 | 1.053 |
| **5** | **PTPN11** | c.1507G>A(p.Gly503Arg) | Absent | Pathogenic | 0.005 | 1.000 | 0.988 | 0.992 | 0.994 | 2.106 | 1.000 | 33.0 | 5.13 | 0.079 | 4.953 |
| **6** | **RYR1** | c.10579C>T(p.Pro3527Ser) | 6.20e-07 | Pathogenic | 0.757 | 0.753 | 0.406 | 0.214 | 0.490 | — | 0.148 | 25.8 | 5.77 | 0.134 | -6.497 |
| **7** | **FOXG1** | c.654C>A(p.Tyr218Ter) | Absent | Pathogenic | — | 1.000 | — | — | — | — | — | 41.0 | 2.79 | 0.139 | 3.796 |
| **8** | **UPB1** | c.319G>A(p.Ala107Thr) | 4.58e-05 | — | 0.001 | 1.000 | 0.898 | 0.921 | 0.891 | 0.561 | 0.361 | 28.3 | 5.06 | 0.755 | 0.549 |
| **9** | **ZC3H14** | c.566T>A(p.Leu189His) | Absent | — | 0.000 | 0.999 | 0.541 | 0.419 | 0.635 | 0.488 | 0.995 | 27.8 | 4.79 | 0.329 | 1.900 |
| **10** | **ROBO3** | c.2579_2580del(p.Val860AlafsTer125) | Absent | — | — | — | — | — | — | — | — | — | — | 0.632 | -0.516 |
| **11** | **MED12L** | c.2365A>G(p.Lys789Glu) | 6.84e-07 | — | 0.017 | 0.992 | 0.755 | 0.703 | 0.302 | 0.703 | 0.872 | 26.0 | 5.35 | 0.174 | 4.008 |
| **12** | **CHD3** | c.9delG(p.Ala4GlnfsTer4) | Absent | — | — | — | — | — | — | — | — | — | — | 0.204 | 8.427 |
| **13** | **GNAO1** | c.736G>A(p.Glu246Lys) | 5.58e-06 | Pathogenic | 0.000 | 0.986 | 0.922 | 0.000 | — | — | 0.999 | 32.0 | — | 0.000 | 4.555 |
| **14** | **ARFGEF1** | c.622C>T(p.Arg208Cys) | 8.21e-06 | — | 0.000 | 1.000 | 0.856 | 0.545 | 0.806 | 1.341 | 0.752 | 31.0 | 5.04 | 0.178 | 6.960 |

*AF, allele frequency; AM, AlphaMissense; CADD, Combined Annotation-Dependent Depletion; GERP, Genomic Evolutionary Rate Profiling; gnomAD, Genome Aggregation Database; MetaRNN, Meta-predictor using Recurrent Neural Network; MPC, Missense badness, PolyPhen-2, and Constraint; MT, MutationTaster; MVP, Missense Variant Pathogenicity; oe_lof, observed/expected loss-of-function ratio; REVEL, Rare Exome Variant Ensemble Learner; SIFT, Sorting Intolerant From Tolerant; VUS, variant of uncertain significance.*
