## Supplementary material for "Challenges and perspectives in implementing whole-exome sequencing in Algeria: lessons from a fully autonomous in-country cohort": Table 1: Clinical characteristics of the cohort

### Table 1. Clinical characteristics of the study cohort.

*Fourteen unrelated patients referred for whole-exome sequencing on the basis of unexplained neurodevelopmental disorders. Referral context: presenting phenotype at the time of referral. Pre-WES genetic staff orientation: diagnostic hypothesis formulated during the internal multidisciplinary staff meeting prior to sequencing. WES outcome: Diagnostic = pathogenic or likely pathogenic variant identified; VUS = variant of uncertain significance; Incidental finding = clinically relevant variant unrelated to the presenting phenotype.*

| Patient | Age range at referral | Sex | Referral context | Pre-WES genetic staff orientation | HPO codes | Paraclinical findings | Cons. | Family history | WES outcome |
|---|---|---|---|---|---|---|---|---|---|
| **1** | **12–17** | M | Syndromic intellectual disability with epilepsy | Severe syndromic ID, Fragile X negative | HP:0012469, HP:0001263, HP:0010864, HP:0001513, HP:0001251, HP:0000486, HP:0002353 | MRI: normal; EEG: diffuse sharp waves, cortical disorganisation | Yes | No | VUS |
| **2** | **8–11** | F | Severe syndromic ID with regression | Clinically suspected Rett syndrome, MECP2 dup. negative | HP:0000252, HP:0002376, HP:0000733, HP:0002371, HP:0001251, HP:0002186, HP:0001250, HP:0001257, HP:0001332, HP:0002650, HP:0005957, HP:0011198 | MRI: normal; EEG: diffuse sharp waves, background slowing | No | No | Diagnostic |
| **3** | **5–7** | F | Psychomotor delay with ataxia and ocular abnormalities | Clinically suspected ataxia-telangiectasia | HP:0001263, HP:0001260, HP:0002304, HP:0001009, HP:0000957, HP:0002205 | NC | Yes | No | Diagnostic |
| **4** | **2–4** | F | Developmental and epileptic encephalopathy | Severe syndromic ID | HP:0001263, HP:0010864, HP:0000252, HP:0000316, HP:0000348, HP:0002162, HP:0001252, HP:0001344, HP:0000729, HP:0002540, HP:0032792, HP:0003155, HP:0011185 | MRI: normal; EEG: temporal sharp waves, tonic seizures | Yes | No | Diagnostic |
| **5** | **8–11** | M | Syndromic short stature with cardiac defect | Clinically suspected Noonan syndrome | HP:0004322, HP:0001508, HP:0000337, HP:0000465, HP:0001642, HP:0011342, HP:0000767, HP:0000316, HP:0000494, HP:0000508, HP:0000369, HP:0000358, HP:0005280, HP:0012810 | MRI: normal; abdominal/pelvic US: normal | No | No | Diagnostic |
| **6** | **5–7** | F | ASD with hearing loss and dysmorphic features | Syndromic ID | HP:0001319, HP:0000729, HP:0001344, HP:0000407, HP:0000337, HP:0000369, HP:0002162, HP:0000233, HP:0000445 | MRI: normal | Yes | No | Incidental finding |
| **7** | **5–7** | F | Global developmental delay with hypotonia | Syndromic ID | HP:0001263, HP:0001252, HP:0001250, HP:0000639, HP:0011185 | EEG: epileptiform discharges, occipital | No | No | Diagnostic |
| **8** | **8–11** | F | Progressive psychomotor regression with epilepsy | Neurometabolic disorder suspected | HP:0001319, HP:0007272, HP:0001250, HP:0002474, HP:0000238, HP:0012448 | MRI: delayed myelination | Yes | No | VUS |

| | | | | | | | | | |
|---|---|---|---|---|---|---|---|---|---|
| **9** | **5–7** | M | Intellectual disability with autistic behaviour | Severe syndromic ID, Fragile X negative | HP:0001250, HP:0002474, HP:0000729, HP:0010864, HP:0001319, HP:0007033 | None | No | No | VUS |
| **10** | **5–7** | M | Congenital nystagmus with progressive scoliosis | Syndromic scoliosis | HP:0007979, HP:0008458, HP:0001252, HP:0012110 | MRI: pontocerebellar hypoplasia | Yes | Two sisters affected | Diagnostic |
| **11** | **2–4** | M | Progressive psychomotor regression with dysmorphia | Neurometabolic disorder suspected | HP:0002376, HP:0001510, HP:0040196, HP:0031956, HP:0031964, HP:0000325, HP:0002232, HP:0001762 | Elevated AST (x2.5); MRI: callosal dysgenesis | Yes | No | VUS |
| **12** | **2–4** | F | Intellectual disability with macrocephaly | Syndromic ID | HP:0001249, HP:0001263, HP:0001252, HP:0001332, HP:0004488, HP:0002007, HP:0000431, HP:0000369, HP:0006808, HP:0001273 | MRI: hypomyelination, callosal dysgenesis | No | No | Diagnostic |
| **13** | **12–23 mo** | F | Hyperkinetic movement disorder with developmental delay | Syndromic neurodevelopmental disorder | HP:0001270, HP:0002072, HP:0001332, HP:0001252, HP:0001257 | MRI: normal; EEG: normal; EMG: normal | No | No | Diagnostic |
| **14** | **0–11 mo** | M | Early developmental impairment | Genetic syndrome | HP:0001250, HP:0001263, HP:0001256, HP:0010851 | MRI: normal; EEG: cortical disorganisation, multifocal sharp waves | No | No | VUS |

*ALP, alkaline phosphatase; ASD, autism spectrum disorder; AST, aspartate aminotransferase; Cons., consanguinity; dup., duplication; EEG, electroencephalogram; GDD, global developmental delay; HPO, Human Phenotype Ontology; ID, intellectual disability; mo, months; MRI, magnetic resonance imaging; NC, not contributory; SNHL, sensorineural hearing loss; US, ultrasound; VUS, variant of uncertain significance; WES, whole-exome sequencing.*
